# Signal change of cerebrospinal fluid with eye drops of O-17-labeled saline

**DOI:** 10.64898/2026.02.12.26346215

**Authors:** Mari Miyata, Moyoko Tomiyasu, Yasuka Sahara, Hiroki Tsuchiya, Takamasa Maeda, Nobuhiro Tomoyori, Makoto Kawashima, Riwa Kishimoto, Atsushi Mizota, Kohsuke Kudo, Takayuki Obata

## Abstract

**Purpose:** Aqueous humor drains fluid from the eye not only via the conventional pathway through the trabecular meshwork and Schlemm’s canal, but also within the eye is known to occur via pathways through the posterior chamber and optic nerve to the cerebrospinal fluid (CSF) surrounding the optic nerve. The mechanism is poorly understood, and non-invasive method for evaluation in living humans has not been established. We previously showed that eye drops containing O-17-labeled water (H_2_^17^O) distribute in the anterior chamber but not the vitreous. This study aimed to evaluate the distribution of H_2_^17^O in the CSF along the optic nerve.

**Methods:** Five ophthalmologically normal participants (20-31 years, all females) were selected from a previous prospective study based on ^1^H MR images of the eyes that included the optic nerve. They received eye drops of 10 mol% H_2_^17^O in their right eye. Dynamic image time series was created by normalizing the signal of each ^1^H-T2WI by the pre-drop average signal. Region-of-interest analyses were performed for signal changes in the anterior chamber, vitreous, and CSF.

**Results:** In the quantitative evaluation, the normalized intensity in the anterior chamber and CSF was significantly lower than that in the pre-drop signal (anterior chamber: 0.78 ± 0.07, p < 0.005; CSF: 0.89 ± 0.07, p < 0.05). No distribution was identified in the vitreous. Qualitatively, the distribution of H_2_^17^O in the anterior chamber was detected in all five participants and in the CSF of four participants (80%).

**Conclusion:** H_2_^17^O eye drops were distributed in the anterior chamber and CSF, but not in the vitreous. These findings suggest that the visualization of aqueous humor outflow, not via the Schlemm’s canal, may contribute to ocular fluid homeostasis, including the ocular glymphatic system.

## Introduction

Aqueous humor outflow is involved in maintaining optimal physiological conditions and ocular homeostasis, including the supply of oxygen and nutrients to inner ocular structures, the removal of metabolic waste, and the regulation of ocular pressure.^1,2^ Increased intraocular pressure due to impaired aqueous humor outflow is the major risk factor for glaucoma.^3^ The flow route of the aqueous humor includes the conventional outflow pathway, which accounts for approximately 70–95% of the drainage through the trabecular meshwork into Schlemm’s canal and then into the collector channels before reaching the episcleral venous system.^2,4,5^ Additionally, the uveoscleral outflow pathway is an unconventional outflow from the anterior chamber through the ciliary muscle interstitial spaces. This pathway eventually leads to the cervical lymph nodes via several outflow pathways, including outflow along the supraciliary and suprachoroidal lumens as well as through efferent ducts that cross the sclera and flow into the choroidal vessels.^6,7^ In the posterior chamber, the aqueous humor that enters the vitreous body is thought to exit into the subretinal space via the retinal pigment epithelium (RPE), which constitutes the outermost layer of the retina, and subsequently drains into the highly permeable choriocapillaris.^8-11^ Furthermore, the presence of lymphatic structures within the dura mater surrounding the optic nerve have been identified in humans,^12,13^ and, in mice, this dural lymphatic pathway has been implicated as a route with high specificity for solute clearance from the vitreous humor.^14^ Thus, when the aqueous humor is drained via pathways other than the conventional outflow pathway, the optic nerve, cerebrospinal fluid (CSF), and dura mater surrounding the optic nerve are presumed to be involved. However, these studies were performed in animal models or *ex vivo*; an *in vivo* method for evaluating the aqueous outflow pathway in humans has not yet been established.

Ideally, MRI can be used to visualize aqueous humor outflow using topical eye drop solutions. Previous MRI studies in humans and animals have demonstrated that eye drops containing deuterium oxide,^15^ gadolinium-based contrast agents,^16^ and oxygen-17 labeled water (H_2_^17^O)^17-19^ can accumulate in the anterior chamber. Among these agents administered via eye drops, only H_2_^17^O is minimally invasive and may be suitable for clinical applications. The chemical exchange of protons between H_2_^17^O and H_2_^16^O leads to the shortening of the T2 relaxation of the H_2_^16^O protons. H_2_^17^O acts as a T2 shortening contrast agent, and its effect can be suppressed by decoupling at the ^17^O frequency during the TE interval of a spin-echo MR sequence.^20^ Consequently, the T2-weighted (T2W) proton MRI (^1^H-MRI) signal intensity in regions containing H_2_^17^O appears lower than in regions containing only H_2_^16^O. Several in vivo studies using dynamic T2W ^1^H-MRI (dT2W_MRI) with H_2_^17^O have been conducted,^17,21,22^ and more recent *in vivo* experiments on the human brain parenchyma and CSF after intravenous administration of 10% or 20% H_2_^17^O saline (1 mL/kg body weight) have been reported.^22,23^

It has been reported that H_2_^17^O saline solution administered as eye drops accumulates in the anterior chamber but cannot be detected in the vitreous body using ^1^H-MR images. However, no evaluation has been conducted to determine whether this can be identified as a drainage pathway via the CSF around the optic nerve. The purpose of this study was to evaluate the physiological distribution of H_2_^17^O in the CSF along the optic nerve in healthy individuals.

## Materials and Methods

### Approval

The Institutional Review Board of the National Institute for Quantum Science and Technology (Chiba, Japan) approved this study (Clinical Trial No. jRCTs031210082). All the procedures were conducted in accordance with the Declaration of Helsinki. Prior to participation, all participants were fully informed about the study, and written informed consent was obtained.

### Study participants

Datasets were acquired from healthy participants, as described by Tomiyasu et al.^18^ Of these six subjects, one was excluded because the CSF surrounding the optic nerve was located outside the field of view. Thus, five participants with no abnormalities upon ophthalmic examination underwent MRI between June and September 2021 (age range 20–37 years, all female).

### Image Acquisition

The MRI protocol and signal quantification of dynamic images were performed as described by Tomiyasu et al.^18^ A 3T MR system (MAGNETOM Verio 3T; Siemens Healthcare, Erlangen, Germany) with a 32-channel head coil was used. The parameters of the dynamic T2W ^1^H-MRI (dT2W_MRI) were as follows: half-Fourier, single-shot, turbo-spin echo sequence,^24^ TR/TE = 3000/444 ms, FOV = 180×180 mm, slice thickness = 3 mm, and matrix = 320×320 pixels, reconstructed to 640×640. The participants started to apply 10 mol% H_2_^17^O saline drops (Taiyo Nippon Sanso, Tokyo, Japan) to their right eye and the number of drops was 20, with a total volume ranging from 0.92 to 1.32 mL. Although the previous protocol acquired images for a longer duration),^18^ the present study analyzed signal intensities from 1 minute before to 9.5 minutes after administration to evaluate early signal changes in the distribution of H_2_^17^O. No images acquired during the period of eye closure were used for data analysis.

### Quantitative and Qualitative assessments

Imaging assessments were independently reviewed by two board-certified neuroradiologists with 30 and 12 years of experience, neither of whom was involved in image processing. For quantitative analysis, the dynamic images were normalized by dividing them by the signal before the administration of H_2_^17^O saline drops as follows:

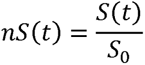

where nS(t) and S(t) are the normalized and original signals at time t, respectively, and S_0_ is the average signal before the application of eye drops for each pixel. The regions of interest (ROIs) were set in the anterior chamber, vitreous body, and CSF along the optic nerve to compare the inflow and outflow rates of H_2_^17^O (Fig. 1).

**Fig. 1.**
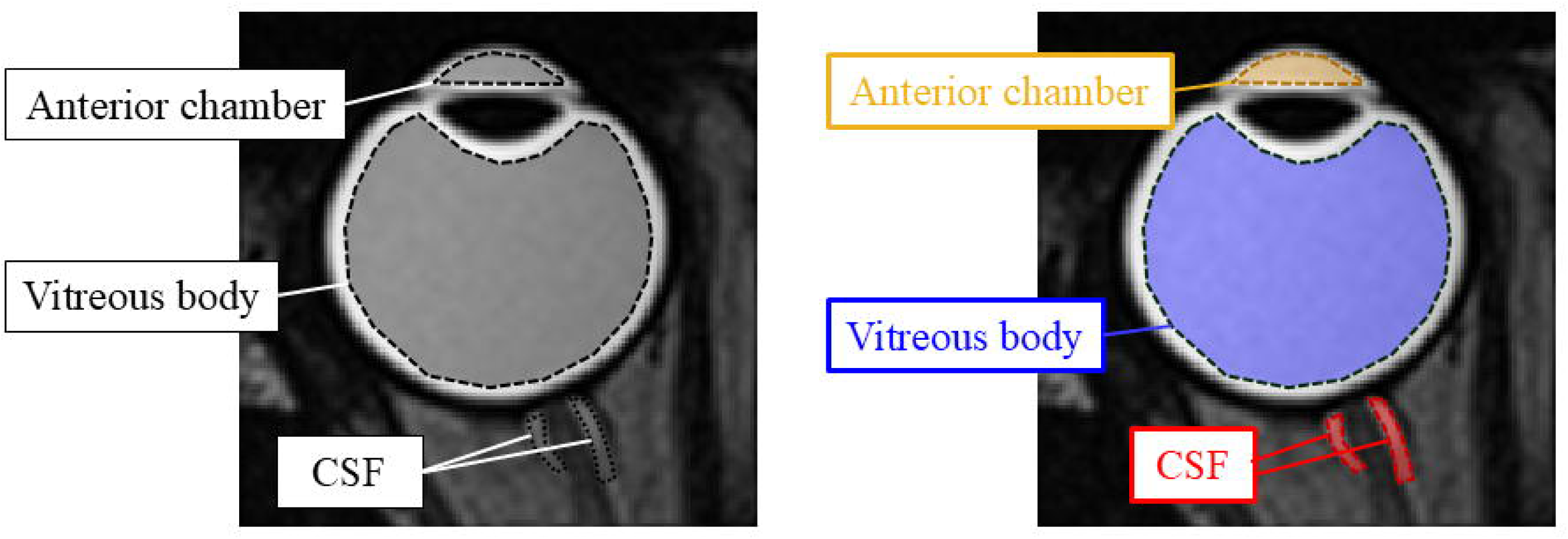
Regions of interest for quantitative evaluation in the right eye.

Three averaged images were used for subsequent qualitative analysis. Each average image was created using 10 images from approximately 5.5, 7, or 8.5 minutes after administering the H_2_^17^O saline drops, excluding images with severe motion artifacts. Finally, subtracted images were obtained by subtracting each average image from the image acquired before the eye drop application. Neuroradiologists independently reviewed these images to determine the presence or absence of H_2_^17^O in the CSF.

### Statistical Analysis

For the image analysis of the signal intensity ratio of the ROI, the average, standard deviation (SD), and range were calculated for the anterior chamber, vitreous body, and CSF along the optic nerve. A paired t-test was used to compare the signal before and after (300-360 seconds) the administration of H_2_^17^O drops in each ROI. A p-value of < 0.05 was regarded as statistically significant. Interobserver agreement for the quantitative assessment was determined by the intraclass correlation coefficient (ICC), using a two-way random-effects model for absolute agreement and single-rater evaluation. ICC values were interpreted as follows: < 0.50, poor reliability; 0.50-0.75, moderate reliability; 0.75-0.9, good reliability; and > 0.90, excellent reliability.^25^ All statistical analyses were performed using MATLAB (MathWorks, Natick, MA, USA).

## Results

According to the qualitative evaluation by the two radiologists, the post-drop normalized signal intensity in the anterior chamber and CSF ROI was significantly lower than the pre-drop intensity for Reader 1 (the anterior chamber: 0.78 ± 0.07, p < 0.005; CSF, 0.89 ± 0.07, p < 0.05), No statistically significant difference was found in the vitreous (1.01 ± 0.03, p = 0.37) (Fig. 2). (The data for Reader 2 are shown in the Supplemental Data.) The ICC between readers for the ROIs was 0.99 (p < 0.0001), demonstrating excellent inter-observer agreement.

**Fig. 2.**
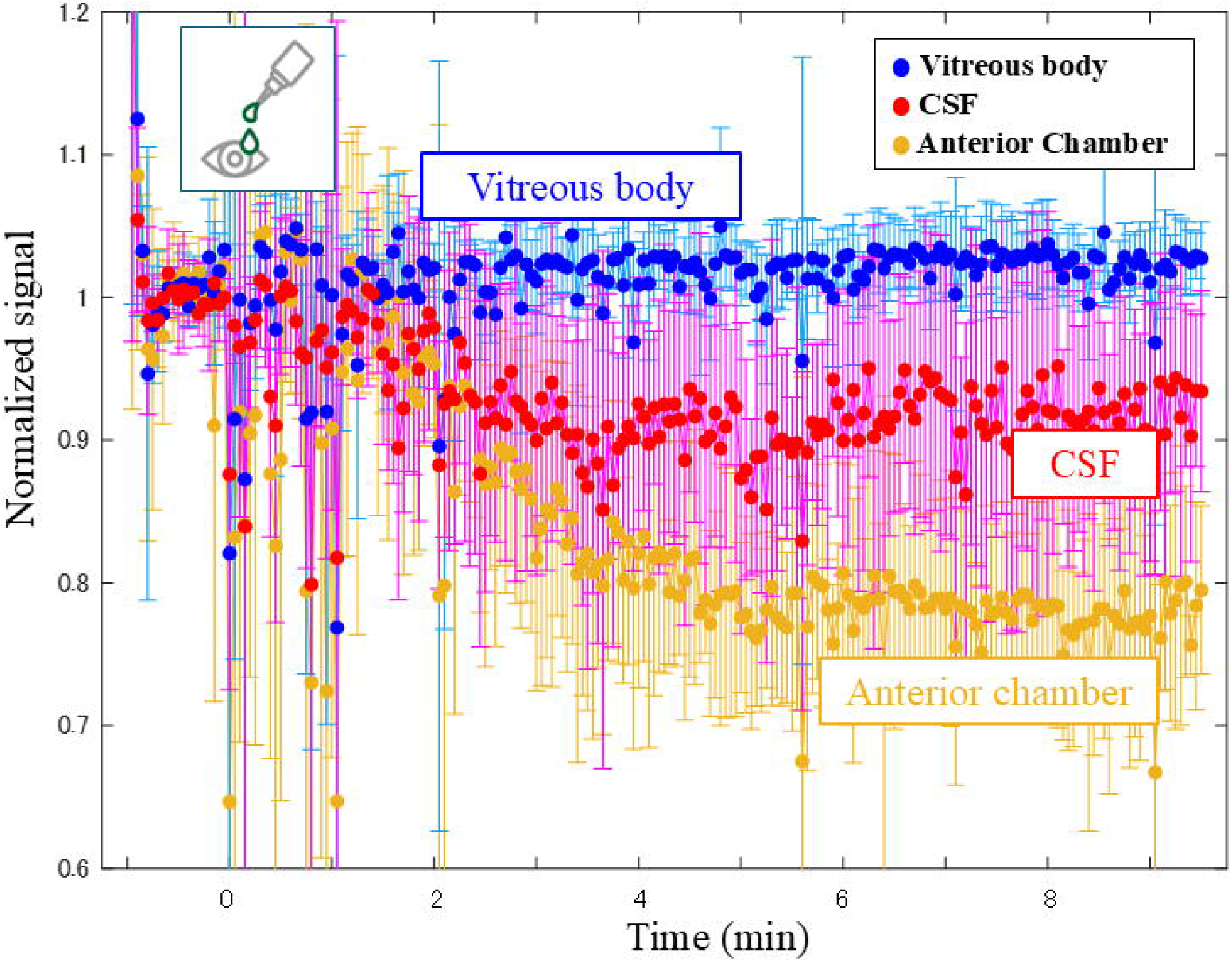
Quantitative evaluation by Reader 1 of time-dependent changes in normalized signal intensity for each region of interest located in the anterior chamber, vitreous body, and cerebrospinal fluid along the optic nerve.

For qualitative evaluation, low signal intensity in the CSF was observed in 4 out of five participants (80%). All participants exhibited low signal intensity in the anterior chamber, but not in the vitreous body (Fig. 3). The results of the qualitative assessments were consistent between the two readers.

**Fig. 3.**
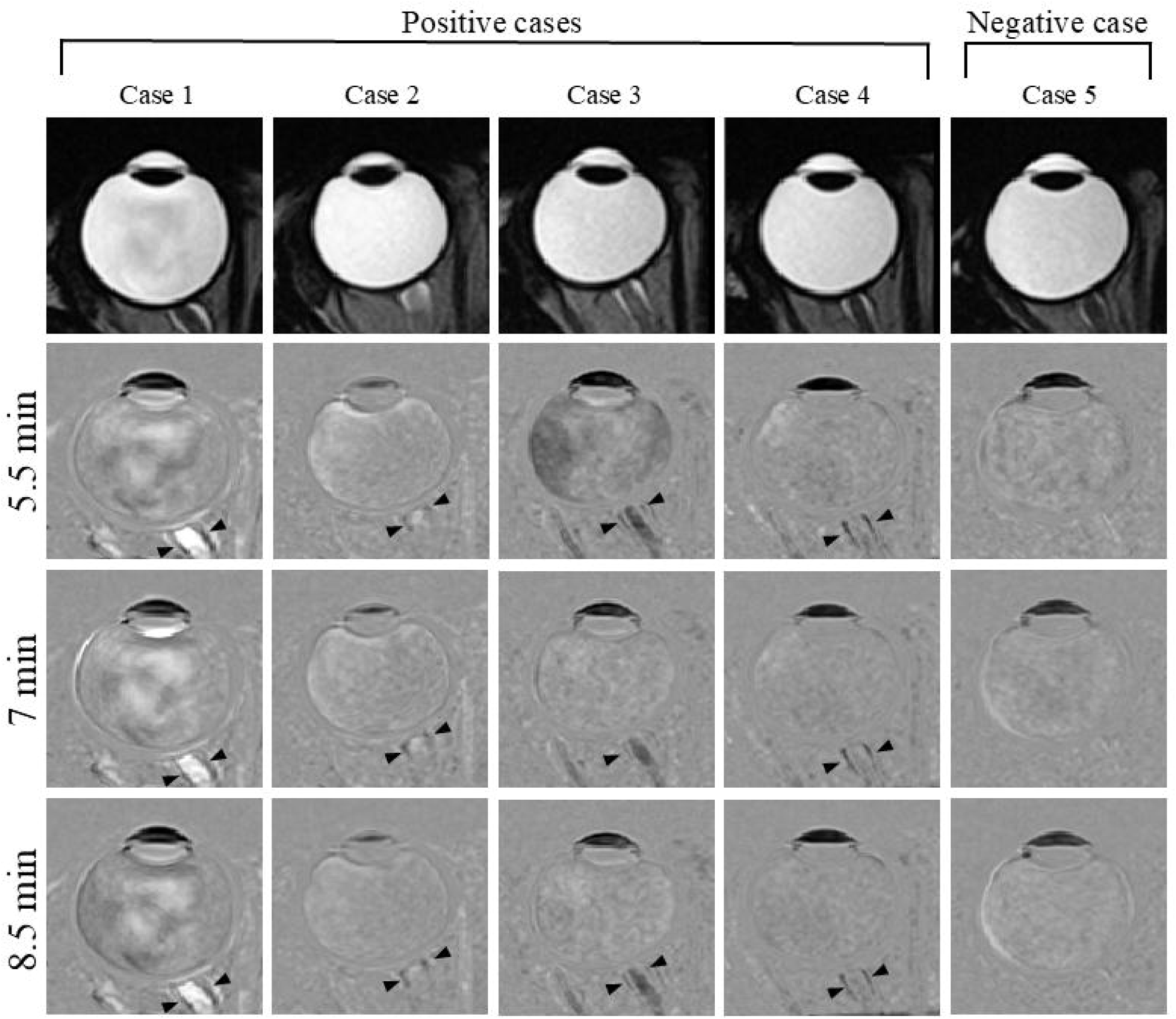
Qualitative evaluation in five participants. Cases 1–4 demonstrated the distribution of H_¹_O in the cerebrospinal fluid along the optic nerve (arrowhead), whereas case 5 showed negative finding.

## Discussion

The present study demonstrated the distribution of H_2_^17^O eye drops in the anterior chamber and CSF along the optic nerve, but not in the vitreous body. To the best of our knowledge, this is the first study to detect aqueous humor outflow *in vivo* by using standard clinical MRI. Previous evaluations of aqueous outflow channels have been limited to animal and ex vivo studies.^26-28^ Thus, this method represents the first minimally invasive and cost-effective technique for the *in vivo* assessment of the human aqueous outflow pathway with potential for clinical implementation.

Previous *in vivo* studies using dT2W_MRI of H_2_^17^O have been conducted in animal models and humans.^17,21,22^ More recently, *in vivo* studies have demonstrated the visualization of H_2_^17^O kinetics in the human brain parenchyma and CSF following intravenous administration of 10% or 20% H_2_^17^O saline (1 mL/kg body weight).^22,23^ Our previous report confirmed the safety of H_2_^17^O eye drops.^18^ The establishment of this novel contrast technique suggests the potential for the clinical implementation of H_2_^17^O imaging, which has been limited by its high cost. Conventional imaging of aqueous humor outflow has been restricted to animal and *ex vivo* models. In contrast, our new H_2_^17^O contrast technique enables the visualization of aqueous humor outflow from the eye to the CSF *in vivo*. Kato et al. previously demonstrated in *ex vivo* pig eyes, the distribution of H_2_^17^O not only in the anterior chamber but also in the vitreous body on dT2W_MRI.^19^ Furthermore, Naganawa et al. suggested that elderly participants who underwent 3T MRI 4 h after standard-dose GBCA administration showed an anteroposterior distribution of GBCA in the vitreous.^29^ However, our results showed that H_2_^17^O was not detected in the vitreous body, but was present in the CSF along the optic nerve. There are several possible reasons for this observation. A similar experiment with rabbits showed distribution in the vitreous body after heavy water drops,^30^ indicating that H_2_^17^O distribution in animal eyes may differ from that in humans. In the human GBCA study by Naganawa et al., participants were relatively old, and the measurement timing was later than that of our study. This indicates that water in the aqueous chamber and CSF may be exchanged through the uvea rather than through the vitreous during the early scan phase in younger individuals. Therefore, this contrast technique, which enables the visualization of intraocular water clearance, may be useful for assessing the pharmacokinetics of eye drops and evaluating aqueous humor dynamics in glaucoma.

Recently, there has been growing interest in the glymphatic system and its essential role in maintaining cerebral homeostasis, particularly through the clearance of metabolic waste. The glymphatic pathway promotes the movement of interstitial fluid (ISF), facilitating the drainage of water and metabolic byproducts such as amyloid-β (Aβ) from the brain into peripheral circulation.^31^ The CSF from the subarachnoid space enters the perivascular pathways ensheathed by astrocytic endfeet,^32^ where water is exchanged between the CSF and ISF via aquaporin-4 (AQP4) channels.

Through these channels, AQP4 also contributes to the elimination of Aβ^31,33^ and hyperphosphorylated tau^34^ both of which are key pathogenic proteins in neurodegenerative diseases. The retina, as the most metabolically active component of the eye, requires continuous removal of neurotoxic waste.^35,36^ However, similar to the brain, the neuroretina and optic nerve lack conventional lymphatic vessels, which are typically responsible for eliminating metabolic byproducts and excess fluid in other tissues. The mechanisms underlying metabolic waste clearance and fluid regulation within the inner retina, particularly in regions with highly active ganglion cells, remain unclear. Following the discovery of the brain glymphatic system,^31^ a corresponding “ocular glymphatic system” has been proposed.^37-41^

Previous reports indicate that following intravitreal administration, Aβ is internalized by retinal ganglion cells and conveyed intra-axonally across the lamina barrier. In contrast, dextran tracers are normally excluded from the optic nerve and gain entry only when the integrity of the lamina barrier is disrupted. Once within the optic nerve, Aβ is released from axons and tends to accumulate along perivenous pathways before being eliminated through the dural lymphatic drainage system.^13,42-44^ The retrograde ocular glymphatic transport process describes the movement of CSF entering the optic nerve along the periarterial routes. From this site, CSF tracers are drained via the dural lymphatic system and subsequently cleared through the cervical lymphatic vessels. Although macromolecular CSF tracers such as dextrans and ovalbumin can reach the lamina barrier, they are unable to penetrate the inner ocular structures.^44,45^ These findings suggested a mechanism analogous to that of the glymphatic system in the brain. In this study, although no signal changes were observed in the vitreous after H_2_^17^O eye drops, distribution in the CSF was detected. This suggests that our new contrast technique may reflect the ocular glymphatic activity *in vivo*.

The present study had several limitations. Firstly, this was a pilot study involving only a small number of participants, who were limited to young females. Future studies include a broader age range and both males and females. Second, because dT2WI was acquired using a single slice, it was impossible to assess whether the eyeball and CSF surrounding the optic nerve were on the same plane, which was the reason for the exclusion of one case. The acquisition of images using 3D dT2WI could overcome this limitation. Third, this study evaluated H_2_^17^O using an indirect method with ^1^H-MRI, although direct methods may yield more accurate quantitative results. Fourth, in the qualitative evaluation, a low CSF signal after H_2_^17^O eye drops was not clearly visible in one participant; this participant had received the lowest dose of eye drops. A detailed assessment of the dosage required to maintain image quality is likely necessary.

## Conclusion

We demonstrated that H_2_^17^O eye drops were distributed in the anterior chamber and CSF, but not in the vitreous body. This novel contrast technique is minimally invasive and cost-effective for clinical implementation, suggesting that the visualization of outflow channels and their lymphatic association may contribute to understanding ocular fluid homeostasis, including the ocular glymphatic system.

## Data Availability

All data produced in the present study are available upon reasonable request to the authors

## Acknowledgment

We thank Kazuko Suzuki and Shizuko Kawakami for their assistance as clinical coordinators.

