## supplemental data for "Signal change of cerebrospinal fluid with eye drops of O-17-labeled saline"

Fig. 1

Quantitative evaluation by Reader 2 of time-dependent changes in normalized signal intensity for each region of interest located in the anterior chamber, vitreous body, and cerebrospinal fluid along the optic nerve. The signal intensity ratio relative to the pre-drop baseline (1 minute starting 5 minutes after eye drop) in the anterior chamber and CSF is significantly lower than that in the pre-drop signal (the anterior chamber: 0.78 ± 0.07, p < 0.005； CSF: 0.90 ± 0.08, p < 0.05). No evaluation identified distribution in the vitreous body (1.01 ± 0.03, p = 0.36).

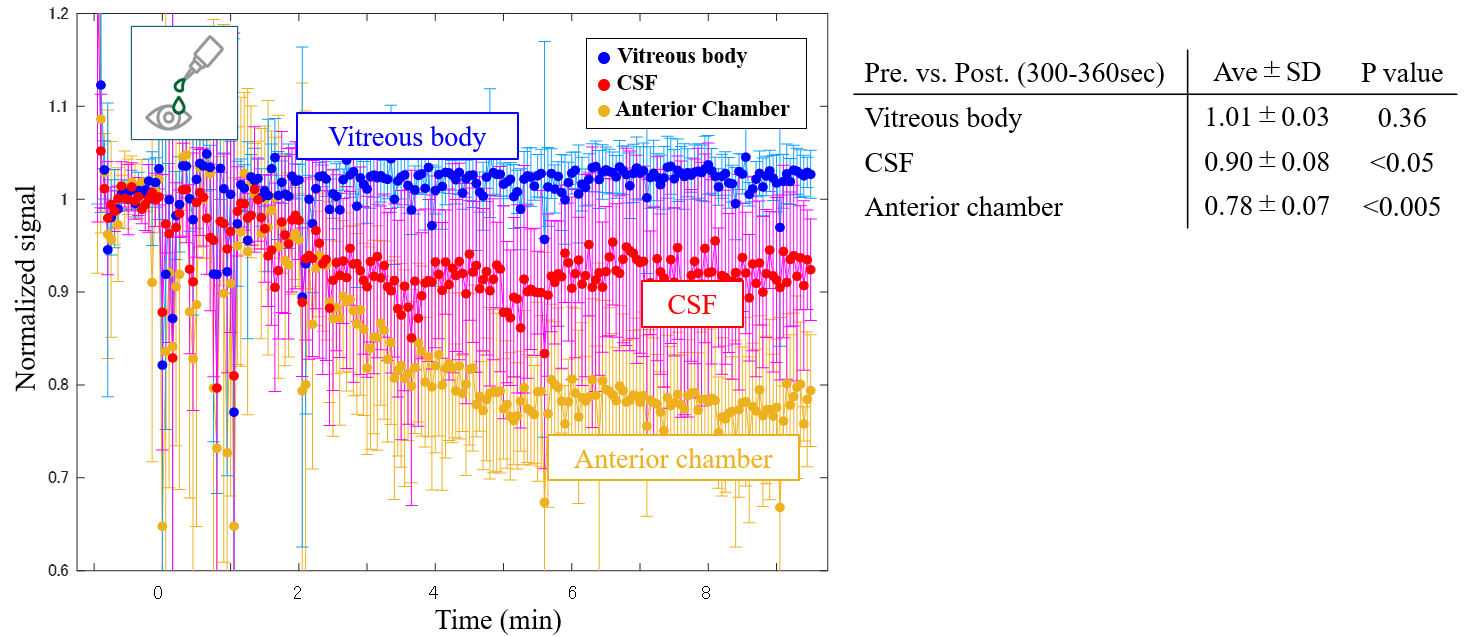
